# Augmented Pain-Evoked Primary Sensorimotor Cortex Activation in Adolescent Girls with Juvenile Fibromyalgia

**DOI:** 10.1101/2022.07.13.22277562

**Authors:** Han Tong, Thomas C. Maloney, Michael F. Payne, Maria Suñol, Christopher D. King, Tracy V. Ting, Susmita Kashikar-Zuck, Robert C. Coghill, Marina López-Solà

## Abstract

**Objective:** Juvenile fibromyalgia (JFM) is a chronic widespread pain condition that primarily affects adolescent girls. Previous studies have found increased sensitivity to noxious pressure in adolescents with JFM. However, the underlying changes in brain systems remain unclear. The aim of this study was to characterize pain-evoked brain responses and identify brain mediators of pain hypersensitivity in adolescent girls with JFM.

**Methods:** Thirty-three adolescent girls with JFM and thirty-three healthy adolescent girls underwent functional MRI scans involving noxious pressure applied to the left thumbnail at an intensity of 2.5 or 4 kg/cm^2^ and rated pain intensity and unpleasantness on a computerized visual analogue scale. We conducted standard general linear model analyses and exploratory whole-brain mediation analyses, and computed pain-evoked brain responses within seven major cortical networks.

**Results:** The JFM group reported significantly greater pain intensity and unpleasantness than the control group in response to noxious pressure stimuli at both intensities (p<0.05). The JFM group showed augmented right primary somatosensory cortex (S1) activation to 4 kg/cm^2^ (Z>3.1, cluster-corrected p<0.05), and the peak S1 activation magnitudes correlated with Widespread Pain Index scores (r=0.35, p=0.048). In the JFM group, we also found augmented activation of the somatomotor network in response to 2.5 kg/cm^2^, and greater primary sensorimotor cortex activation in response to 4kg/cm^2^ mediated the between-group differences in pain intensity ratings (p<0.001).

**Conclusion:** We found heightened sensitivity to noxious pressure stimuli and augmented pain-evoked sensorimotor cortex responses in adolescent girls with JFM, which could reflect central sensitization or amplified nociceptive input.

## Introduction

Juvenile-onset fibromyalgia (JFM) is a poorly understood chronic pain condition that affects up to 6% of children, primarily adolescent girls(1). It is characterized by persistent widespread musculoskeletal pain and often accompanied by physical fatigue, nonrestorative sleep, headaches, anxiety, and depression(2). Unfortunately, there is no known cure for JFM, and over 80% of adolescents with JFM continue to experience pain into adulthood(3). Despite sharing many clinical features(4), important differences exist between JFM and adult-onset fibromyalgia (FM). First, joint hypermobility and dysautonomia, rarely reported in adults with FM, are common in adolescents with JFM(2, 5). Secondly, the brain, which is critical for forming and regulating pain perception(6), undergoes robust development during adolescence(7). In a recent article, we were the first to report that brain processing of pain during adolescence differs from adulthood(8), suggesting that the conclusions from adult fibromyalgia studies may not be directly applicable to JFM pathophysiology.

Although the past two decades have seen considerable advances in understanding the brain mechanisms of adult fibromyalgia(9-15), similar mechanistic research on JFM is mostly lacking. In a 2017 study using quantitative sensory testing, our group observed that adolescents with JFM were more sensitive to noxious pressure than their healthy peers(16), suggesting a propensity for sensitization of peripheral and/or central nociception. In our group’s recent structural MRI study, adolescents with JFM showed significantly less grey matter volume in the pain-associated midcingulate cortex than healthy controls(17). To our knowledge, only one preliminary study examined brain activity related to pressure pain in adolescents with JFM(18). However, this study was likely limited by the small sample size (only 10 participants per group) and uncorrected statistics. The aim of the present study was to investigate the brain correlates of the observed pain hypersensitivity in adolescents with JFM. Using a functional magnetic resonance imaging (fMRI) task involving noxious pressure stimulation, we compared pain-evoked brain responses between 33 adolescent girls with JFM and 33 healthy adolescent girls. We conducted standard massive univariate general linear model analyses, computed pain-evoked brain activation responses within major cortical brain networks and performed exploratory whole-brain mediation analyses. Previous fMRI studies show that adult fibromyalgia is associated with augmented brain responses to acute experimental pain involving primary somatosensory cortex, secondary somatosensory cortex, posterior and anterior insula, and anterior mid-cingulate cortex(9-12, 14, 15). Based on these findings, we anticipate that adolescents with JFM may also exhibit significantly augmented pain-evoked brain responses in one or more of these pain-processing regions(6) previously identified in adults.

## Patients and Methods

### Participants

This study involved 33 adolescent girls with JFM (13-18 years old, mean age of 15.85 ± 1.09 years) and 33 matched healthy girls (13-18 years old, mean age of 15.33 ± 1.31 years; two-sample t-test for age, t=1.73, p=0.089). Inclusion and exclusion criteria for study participants are detailed in Supplementary Material. Before being enrolled in the study, legal guardians of the study participants provided written informed consent and all participants provided informed assent. The study protocol and consent forms were approved by Cincinnati Children’s Hospital Medical Center Institutional Review Board (Study ID: 2017-7771). All of the data for this study were collected between February 2018 and March 2021.

### Study procedures

This study is part of a larger ongoing clinical study on brain mechanisms of juvenile fibromyalgia which has been registered on ClinicalTrials.gov (Study ID: NCT03612258).

All participants completed two study visits. Visit 1 involved collecting demographic information, administering clinical questionnaires and familiarizing the participants with the pressure pain fMRI task. Clinical questionnaires included Widespread Pain Index which assesses the number of pain locations, Symptom Severity Scale which assesses the severity of three cardinal symptoms of fibromyalgia (i.e., fatigue, waking unrefreshed, cognitive symptoms) and the presence of other somatic symptoms, as described in the 2010 ACR diagnostic criteria for FM(4, 19). These are two subscales from the Pain and Symptom Assessment Tool (PSAT), which was developed to enhance consistent classification of adolescents with juvenile fibromyalgia (JFM) based on the 2010 ACR diagnostic criteria for FM(20).

During Visit 1, the experimenter explained the pressure pain task and demonstrated the pressure stimulation device to the participants. Then, participants were asked to practice the pain rating task on a computer. Noxious pressure was applied with a hand-held algometer using the same stimulus intensity, duration, and interval as the stimuli administered by the pressure pain device during the fMRI task.

Visit 2 immediately followed Visit 1 and involved MRI data acquisition. During the pressure pain fMRI task, a computer-controlled pneumatic device, which could reliably transmit preset pressure to 1-cm^2^ surface (8, 9, 11, 12, 21), was used to deliver noxious pressure stimuli to the left thumbnail. Noxious pressure was applied at the intensity of either 2.5 kg/cm^2^ or 4 kg/cm^2^.

A block design was adopted for our fMRI task, which was programmed and presented to the participants using the E-Prime 3.0 software (Psychology Software Tools, Pittsburgh, PA). The task consisted of two consecutive fMRI runs, each containing 6 trials in a mixed pseudorandom order, with 3 trials at each stimulus intensity. As shown in **Figure S1**, each trial began with a rest period with pseudorandom duration (range: 11-20 seconds), followed by a very brief auditory cue lasting 0.2 seconds, a 3-to-6-second pain anticipation period, and then a fixed 10-second pain period. After an 8-10 second post-pain period, the participants were asked to rate the pain intensity (“How intense was the pain you just experienced?”) and pain unpleasantness (“How unpleasant was the pain you just experienced?”) on computerized visual analogue scales (VAS) from 0 (“not painful/unpleasant at all”) to 100 (“most painful/unpleasant imaginable”)(22). The participants were instructed to move the cursor on the scales using an MRI-compatible trackball until the position that best describes their pain experience and click the button to submit their ratings. The numbers between 0 and 100 on the scales were not visible to the participants. At the end of each run, the participants were asked to rate intensity of spontaneous bodily pain (“How much bodily pain did you experience during the task?”) and extensiveness of spontaneous bodily pain (“How extensive was the bodily pain you felt during the task?”) unrelated to the noxious stimuli on a computerized VAS from 0 (“no pain at all” or “very localized pain”) to 100(“worst pain imaginable” or “pain all over my body”). Answers to these two questions represented the self-reported severity of spontaneous bodily pain during the task.

### Magnetic resonance imaging data acquisition

Neuroimaging data for this study were acquired at Cincinnati Children’s Hospital Medical Center using a Philips Ingenia 3T MR System (Philips Healthcare, Best, The Netherlands) with a 32-channel head coil. Structural MRI images of the brain were acquired using the standard T1 weighted gradient echo sequence and the following scan parameters: TR = 10 ms, TE = 1.8, 3.8, 5.8, 7.8 ms, field of view = 256 × 224 × 200 mm, voxel size = 1 × 1 × 1 mm, number of slices = 200, flip angle = 8°, slice orientation = sagittal, and scan duration = 4:42 minutes. Blood oxygen level-dependent (BOLD) fMRI data were collected using T2* weighted echo planar imaging sequence with multiband sensitivity encoding (SENSE) technique(23). Scan parameters for the pressure pain task were as follows: multiband acceleration factor = 4, TR = 650 ms, TE=30 ms, field of view = 200 mm, flip angle = 53°, voxel size =2.5 × 2.5 × 3.5 mm, slice orientation = transverse (parallel to the orbitofrontal cortex line), slice thickness = 3.5 mm, number of slices = 40 (provided whole-brain coverage), number of volumes = 522, number of dummy scans = 12, and scan duration = 5:42 minutes. Two consecutive fMRI runs of pressure pain task were acquired. Information regarding other fMRI scans acquired during the same MRI session can be found on ClinicalTrials.gov (Study ID: NCT03612258).

## Data analyses

### Statistical analyses of behavioral data

We built a mixed-design ANOVA model with “group” as the between-subject variable and “stimulus intensity” as the within-subject variable using R software (version 3.6.2, R Foundation for Statistical Computing, Vienna, Austria) to assess differences in pain intensity and unpleasantness between the JFM group and the healthy adolescent group under two experimental conditions (i.e., noxious pressure stimuli at 2.5kg/cm^2^ and 4kg/cm^2^). Post-hoc pairwise comparisons were made using the false discovery rate (FDR) correction method. In addition, we compared the intensity and extensiveness of bodily pain between the two groups using two-sample t-tests and computed Pearson’s correlations between pain ratings to noxious pressure stimuli (2.5 kg/cm^2^ and 4 kg/cm^2^) and bodily pain ratings (intensity and extensiveness), respectively.

### Preprocessing of neuroimaging data

We preprocessed the neuroimaging data using FSL (FMRIB Software Library version 6.0.3, the Analysis Group, FMRIB, Oxford, UK)(24) and AFNI (Analysis of Functional Neuroimages version 20.3.02, Medical College of Wisconsin, WI, USA)(25). For the structural images, we performed brain extraction using Brain Extraction Tool (BET)(26) in FSL. We carried out bias correction and segmentation using FMRIB’s Automated Segmentation Tool (FAST) in FSL(27). Then we normalized and resampled the brain-extracted image to the 2-mm isotropic MNI ICBM 152 non-linear 6th generation template(28) using FSL’s FMRIB’s Linear Image Registration Tool (FLIRT)(29, 30). We preprocessed each participant’s functional scans in the following steps: First, brain extraction was performed using FSL’s BET(26). Next, outlying functional volumes (i.e. spikes) were detected using the DVARS metric within FSL’s “fsl_motion_outliers”(31). We performed motion correction of the functional time-series using MCFLIRT(30). The motion corrected data were then high-pass filtered at 0.00556 Hz (180 seconds) and smoothed with a 6 mm full width at half maximum (FWHM) filter using AFNI’s 3dBandpass. To minimize pain-induced global cerebral blood flow fluctuations(32), we applied intensity normalization by scaling each fMRI volume by its mean global intensity. The intensity-normalized data were first co-registered with the participant’s T1 image using FSL’s FLIRT (6-parameter rigid body model)(30), then aligned to the MNI template(28).

### General linear model analyses

We used the general linear model (GLM) approach as implemented in FSL’s “fsl_glm”(33) to estimate each participant’s brain responses to noxious stimuli. Specifically, we modeled the three pain periods associated with 2.5 kg/cm^2^ stimuli and the other three pain periods associated with 4 kg/cm^2^ stimuli as two separate regressors. In addition to the pain period regressors, our GLM model included regressors for the anticipatory periods, post-pain periods, and pain rating periods.

The remaining “rest” period was used as the implicit baseline. Finally, six motion parameters (3 for translational motion and 3 for rotational motion) and outlying volumes (“spikes”) were included as nuisance regressors. The two runs of each participant’s first-level GLM results, which included estimated contrasts of parameter estimates (COPEs) and their variances (VARCOPEs), were combined at the single-subject level using the fixed effects modeling in FSL with “flameo”(34). Then at the group-level, mixed effects modeling (FLAME 1+2)(34) was used to compute each group’s mean brain responses to pressure pain (one-sample t-test) and between-group differences (two-sample t-test) for each condition (2.5 kg/cm^2^ and 4 kg/cm^2^). The group-level GLM analyses for between-group differences were repeated with age added as a covariate in the model to verify that the observed group difference was not explained by age. The results of group-level analyses were corrected for multiple comparisons across the whole brain using FSL’s “cluster” tool. Clusters of voxels were identified using a threshold of Z>3.1 and their statistical significance (p<0.05) was estimated by cluster-based inference according to Gaussian random field theory(35). The maximum Z-score within the region representing the between-group difference in brain responses to noxious stimuli was extracted for all participants with JFM and correlated with their Widespread Pain Index and Symptom Severity Scale scores.

### Pain-evoked neural responses in large-scale brain networks

To examine pain-evoked neural responses in large-scale brain networks, using NumPy in Python(36), we computed the dot product of each participant’s contrast images of parameter estimates (i.e., “pressure pain at 2.5 kg/cm^2^” and “pressure pain at 4 kg/cm^2^”) for each run and pre-defined masks of the previously identified seven major resting-state cortical networks(37), including the somatomotor network, the fronto-parietal network, the dorsal attentional network, the ventral attentional network, the limbic network, the default mode network, and the visual network. Then, the responses within each brain network for two runs were averaged at the individual participant level, which resulted in a single-scalar value as a summary metric representing overall pain-evoked neural responses across the entire functional brain network. Lastly, between-group comparisons for each network and each condition were made in R software using two-sample t-tests.

### Exploratory whole-brain mediation analyses

First-level contrast images for the pain period regressors were carried forward to a whole-brain mediation analysis model. We tested relationships between group (adolescents with JFM versus healthy adolescents), single-subject pain-evoked brain responses (contrast images for each trial), and single-subject pain intensity ratings to stimuli at 4 kg/cm^2^ using mediation analysis found in the multilevel mediation and moderation (M3) toolbox(38) and implemented in MATLAB (version R2019b, MathWorks, MA, USA)(39-41). Mediation analysis identifies brain regions that show partially independent, but not orthogonal, effects: (1) between-group differences in pain-evoked brain responses (path a), (2) brain activity that predicts changes in pain intensity ratings (path b) controlling for group, and (3) brain activity mediating the between-group differences in pain intensity ratings (path a x b). The resulting activation maps were shown at uncorrected p<0.001, as implemented in our previous study(8). To facilitate interpretation of the functional maps, adjacent voxels to the identified clusters were also displayed in a different color at a lower threshold of uncorrected p<0.005.

## Results

### Adolescents with JFM show greater pain sensitivity to noxious pressure

As shown in **Figure 1**, pain intensity and pain unpleasantness ratings in response to noxious stimuli at 2.5 kg/cm^2^ were 31.64 ± 20.72 (mean ± standard deviation) and 33.72 ± 22.44 in adolescents with JFM, and 20.66 ± 13.12 and 19.31 ± 11.82 in healthy adolescents, respectively. Pain intensity and pain unpleasantness ratings to noxious stimuli at 4 kg/cm^2^ were 41.38 ± 21.07 (mean ± standard deviation) and 44.43 ± 20.86 in adolescents with JFM and 28.49 ± 16.61 and 29.58 ± 17.22 in healthy adolescents, respectively (measured using a VAS ranging from 0 to 100). Using a group-by-stimulus intensity mixed-design ANOVA, we found a significant main effect of group (pain intensity: F=7.48, p=0.008; pain unpleasantness: F=11.16, p=0.001) and a significant main effect of pressure on pain ratings (pain intensity: F=77.97, p<0.001; pain unpleasantness: F=65.96, p<0.001), but did not find an interaction between group and stimulus intensity (pain intensity: F=0.92, p=0.34; pain unpleasantness: F=0.03, p=0.865). Post-hoc pairwise comparisons showed that, in response to stimuli at either 2.5 or 4 kg/cm^2^, adolescents with JFM reported greater pain intensity (2.5 kg/cm^2^: t=2.57, FDR-corrected p=0.013; 4 kg/cm^2^: t=2.76, FDR-corrected p=0.010) and pain unpleasantness (2.5 kg/cm^2^: t=3.26, FDR-corrected p=0.002; 4 kg/cm^2^: t=3.15, FDR-corrected p=0.003) than healthy adolescents. These results indicate that adolescents with JFM have greater pain sensitivity to noxious pressure stimuli than their healthy peers for both intensities of noxious stimuli.

**Figure 1.**
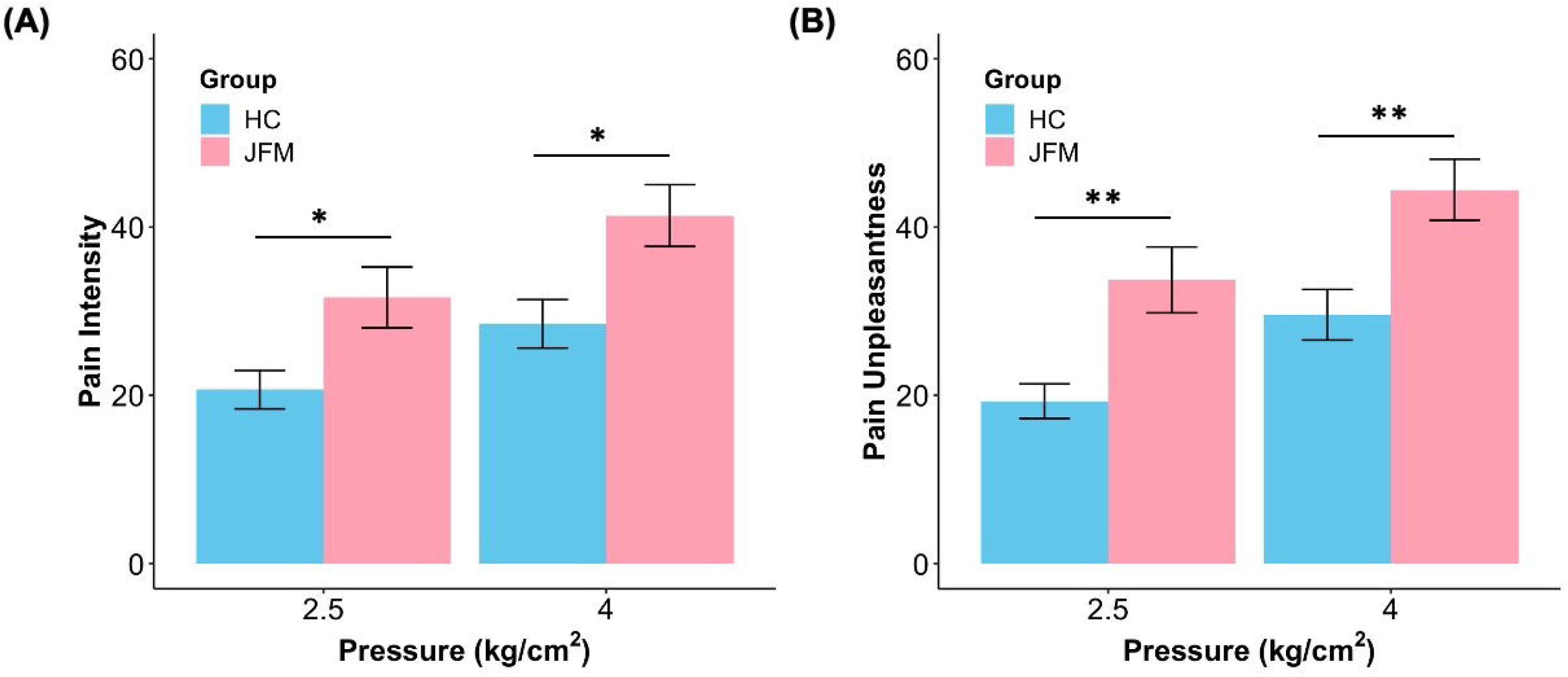
Pain intensity (A) and unpleasantness (B) ratings to noxious pressure stimuli by different (2.5, 4 kg/cm^2^) pressure in healthy adolescents (blue bar) and adolescents with JFM (pink bar). Using mixed-design ANOVA tests, we found a significant main effect of group and a significant main effect of pressure on pain ratings but did not find an interaction between group and stimulus intensity neither for pain intensity nor for pain unpleasantness. *p<0.05 in post-hoc tests (FDR corrected); **p<0.01 in post-hoc test (FDR corrected).

### Experimental pain ratings and spontaneous bodily pain ratings are significantly correlated in adolescents with JFM

As expected, adolescents with JFM reported significantly greater intensity (t=9.42, p<0.001) and extensiveness (t=8.04, p<0.001) of spontaneous bodily pain during the task (i.e., pain unrelated to the noxious stimuli) than healthy adolescents (**Figure S2**).

In adolescents with JFM, average pain intensity ratings to 2.5kg/cm^2^ (a total of 6 trials) were positively correlated with the intensity (t=3.61, p=0.001, r=0.54) and extensiveness (t=2.29, p=0.029, r=0.38) of spontaneous bodily pain (unrelated to the noxious stimuli) experienced during the fMRI scan and reported at the end of the scan also using a VAS. Average pain intensity ratings to 4 kg/cm^2^ (6 trials) also showed a positive correlation with the intensity of spontaneous bodily pain experienced along the course of the scan (t=2.56, p=0.016, r=0.42) and a trend towards a positive correlation with extensiveness of spontaneous bodily pain (t=1.76, p=0.088, r=0.30) (**Figure 2**).

**Figure 2.**
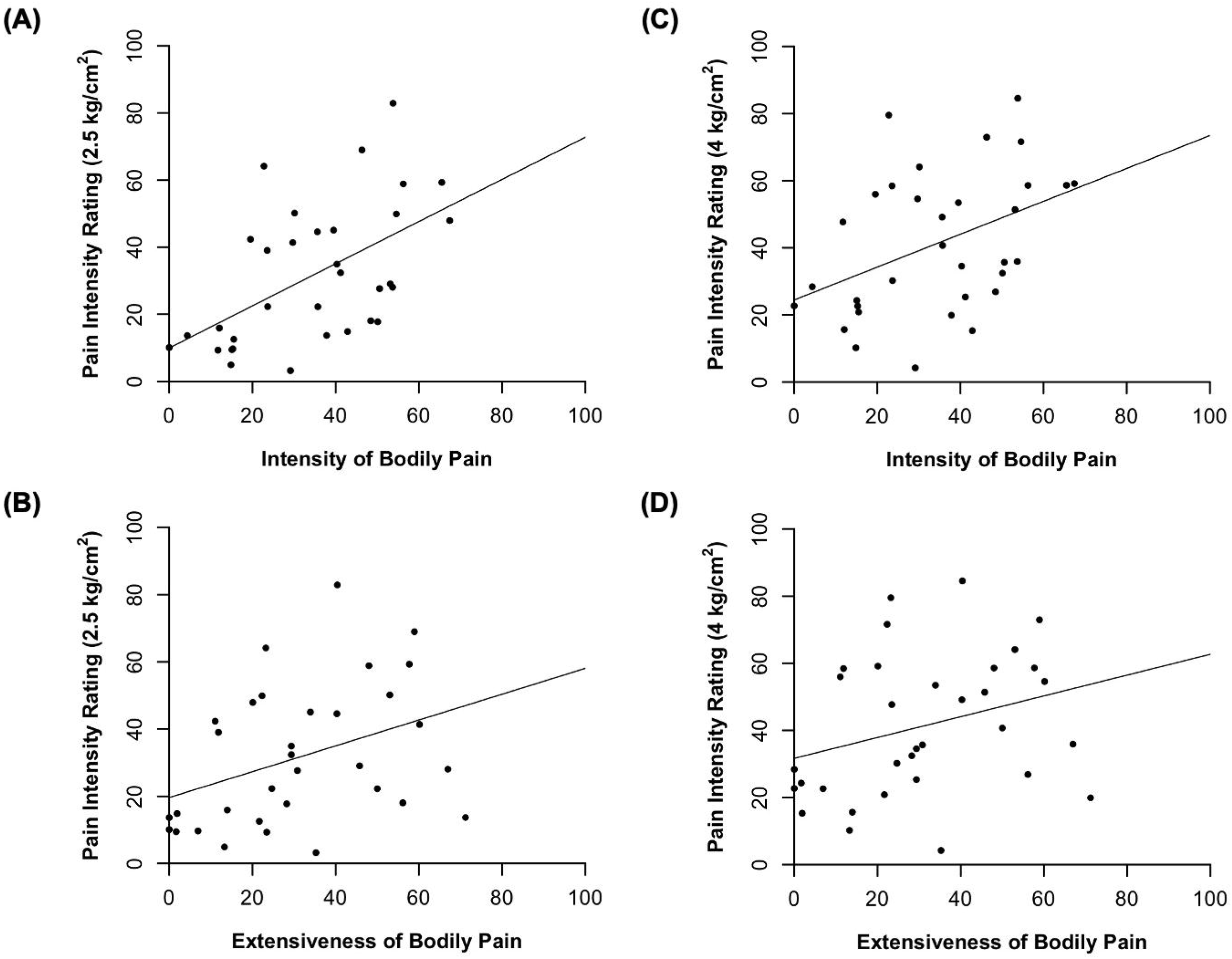
Associations between pressure pain and bodily pain in patients with JFM. Pain intensity ratings to 2.5 kg/cm^2^ (left column) were positively correlated with (A) intensity of bodily pain (t=3.61, p=0.001, r=0.54) and (B) extensiveness of bodily pain (t=2.29, p=0.029, r=0.38). Pain intensity ratings to 4 kg/cm^2^ (right column) also showed a positive correlation with (C) intensity of bodily pain (t=2.56, p=0.016, r=0.42), and a trend towards a positive correlation with (D) extensiveness of bodily pain (t=1.76, p=0.088, r=0.30).

### Adolescents with JFM show augmented pain-evoked activation in primary somatosensory cortex, and activation in such region correlates with widespreadness of spontaneous bodily pain

Adolescents with JFM showed pain-evoked brain activation in regions similar to those found in healthy adolescents, including bilateral insular cortex and operculum, parietal operculum (S2), supramarginal gyrus, primary sensorimotor cortex (S1/M1), anterior cingulate cortex, supplementary motor area, dorsolateral prefrontal cortex, superior temporal gyrus, basal ganglia, thalamus and amygdala (**Figure 3** and **Table S1-S4**). Both groups of adolescents showed pain-evoked deactivations in the fusiform gyrus, precuneus cortex, and occipital visual cortex. In both groups, the cerebellum was deactivated in response to 2.5kg/cm^2^ and became activated in response to stimuli at 4kg/cm^2^. When compared statistically, adolescents with JFM exhibited significantly augmented activation (Z>3.1, p<0.05 corrected for multiple comparisons) in the right primary somatosensory cortex (S1, postcentral gyrus) in response to noxious stimuli at 4 kg/cm^2^ (**Figure 3 and Table S5**). Brain responses to noxious stimuli at 2.5 kg/cm^2^ were not significantly different between groups after correcting for multiple comparisons (**Figure 3**). After adjusting for age, between-group differences in S1 remained significant for stimuli at 4 kg/cm^2^ and became significant for stimuli at 2.5 kg/cm^2^ as well (**Table S6-S7**). Importantly, we found a statistically significant positive correlation between pain-evoked responses in the region of S1 showing the largest between-group effects and the Widespread Pain Index scores in adolescents with JFM (r=0.35, p=0.048, **Figure 4**). In contrast, there was no such association between S1 activity and Symptom Severity Scale scores (p=0.93, r=-0.01).

**Figure 3.**
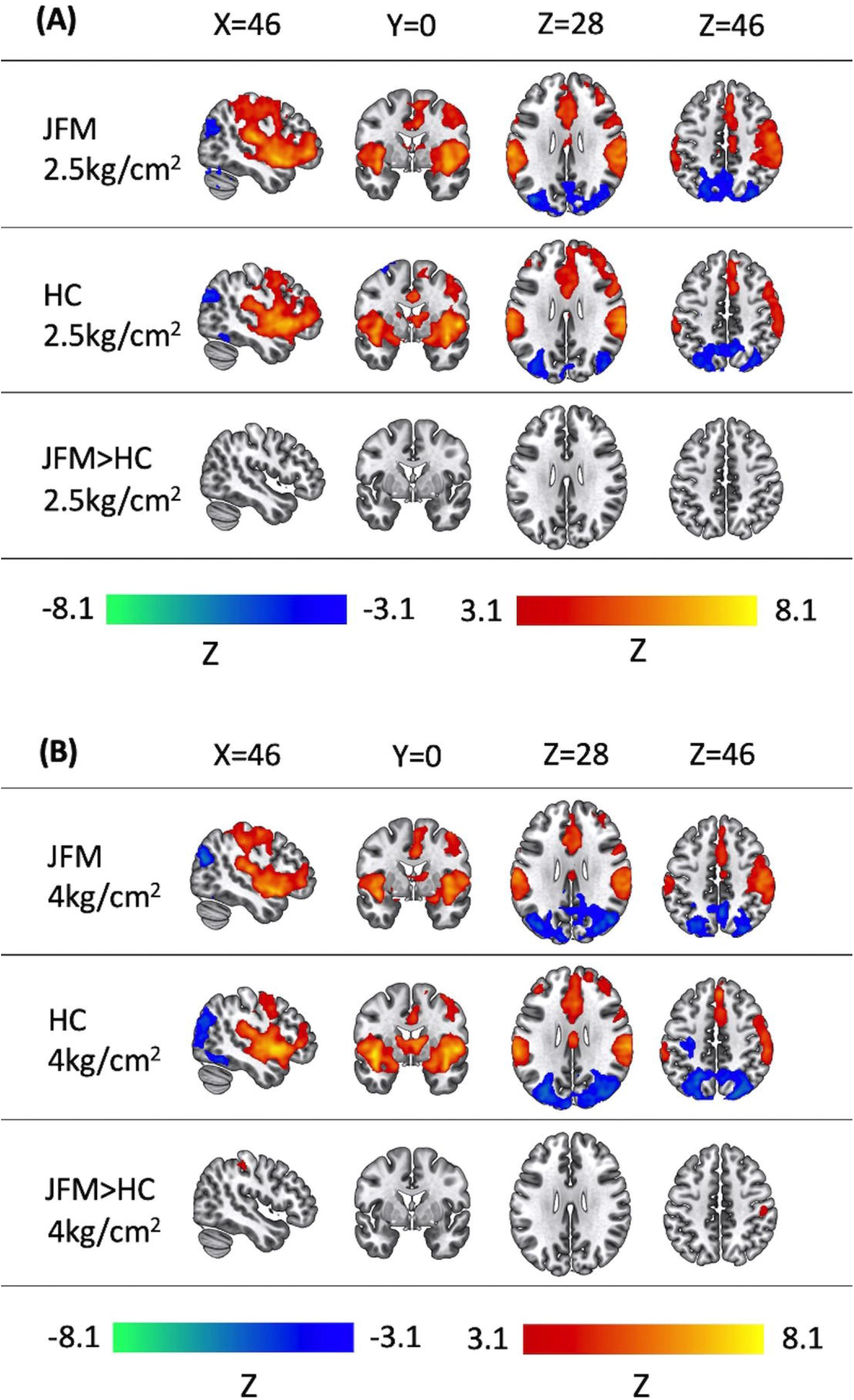
Pain-evoked brain responses in adolescents with JFM (top rows), healthy adolescents (middle rows), and between group comparisons (bottom rows). (A) Brain responses to 2.5 kg/cm^2^ were not significantly different between the groups. (B) Adolescents with JFM had significantly greater activation in right primary somatosensory cortex (postcentral gyrus, S1) in response to stimuli at 4 kg/cm^2^. Statistical significance was estimated using cluster-based inference at Z>3.1, cluster-corrected p<0.05.

**Figure 4.**
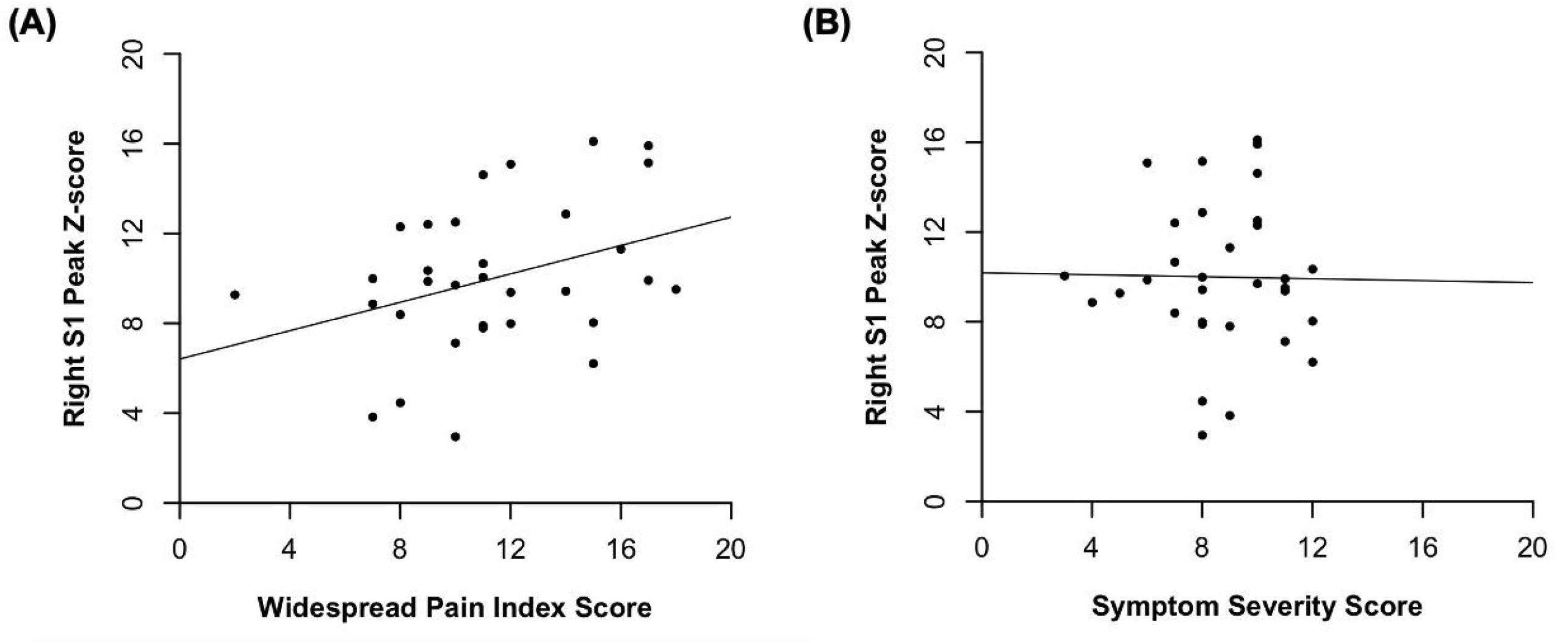
Association of peak pain-evoked activation in right S1 and two Pain and Symptom Assessment Tool subscales in adolescents with JFM. (A) Widespread Pain Index scores were positively correlated with peak activation magnitude (Z-score) within right primary somatosensory cortex (S1), corresponding to the between-group difference in brain responses to 4 kg/cm^2^ stimuli (t=2.06, p=0.048, r=0.35). (B) Symptom Severity scores were not correlated with peak S1 brain activation (t=0.08, p=0.93, r=-0.01).

### Adolescents with JFM show greater pain-related activation in the somatomotor network

In both groups, we found significant pain-related activation in the somatomotor network, the frontoparietal network and the ventral attention network, and pain-related deactivation in the dorsal attention network and the visual network for both stimulus intensities (one-sample t-test, p<0.05). During painful stimulation, the default mode and the limbic networks were neither significantly activated nor deactivated (**Table S8**). Importantly, compared to the healthy control group, the JFM group showed significantly augmented pain-related activation in the somatomotor network in response to stimuli at 2.5 kg/cm^2^ (two-sample t-test, t=2.59, p=0.012). The results for somatomotor network responses to stimuli at 4 kg/cm^2^ showed a similar trend, though the between-group differences were not statistically significant (t=1.51, p=0.136) (**Figure 5**). Brain responses in other networks were not significantly different between the two groups (**Table S9**).

**Figure 5.**
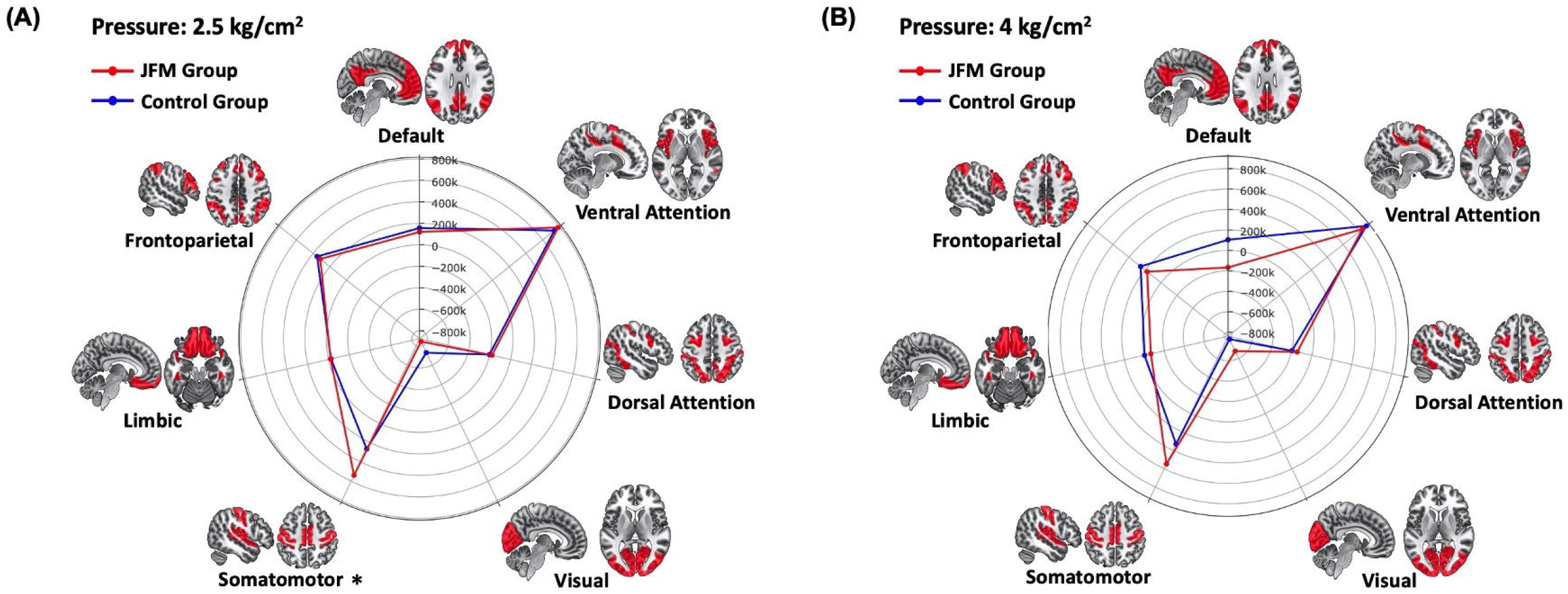
Polar plots comparing pain-evoked brain responses to noxious pressure stimuli at (A) 2.5 kg/cm^2^ and (B) 4 kg/cm^2^ between the JFM group and the control group within 7 major cortical networks (Yeo BT et al., 2011). The numerical values are the group means of the dot product of the pre-defined masks of these networks and each participant’s contrast images of parameter estimates for the pain period (2.5 kg/cm2 or 4 kg/cm2). * p<0.05 in two-sample t-test.

### Augmented pain-evoked sensorimotor activation mediates greater pain sensitivity in adolescents with JFM

We performed an exploratory whole-brain mediation analysis to investigate brain activity underlying the observed group difference in pain intensity ratings between adolescents with JFM and healthy adolescents. As illustrated in **Figure 6**, the mediation model included “group (JFM vs. Control)” as the independent variable X, “pain intensity ratings to stimuli at 4 kg/cm^2^” as the dependent variable Y, and “brain responses to stimuli at 4 kg/cm^2^” as the mediator M. Path a (X→M) represents the between-group differences in brain responses to noxious pressure stimuli. In line with the results of the GLM analyses, we found that adolescents with JFM had greater pain-evoked responses than healthy adolescents in the right primary sensorimotor cortex (precentral and postcentral gyrus), contralateral to the side of body receiving noxious stimuli. A subregion of the left primary sensory cortex and the right superior parietal lobule also showed greater responses in JFM. Path b (M→Y) represents pain-evoked brain activity predicting higher pain intensity ratings controlling for group. Significant path b effects were found in the following regions: cerebellum, thalamus, precuneus, ventromedial prefrontal cortex, and primary sensorimotor cortex. Path a x b (X→M→Y) represents the mediation effect. We found that brain activity in the primary sensorimotor cortex mediated group effects on pain intensity ratings. In other words, these results suggest that augmented pain-evoked sensorimotor responses in adolescents with JFM account for their augmented pain sensitivity compared to healthy adolescents.

**Figure 6.**
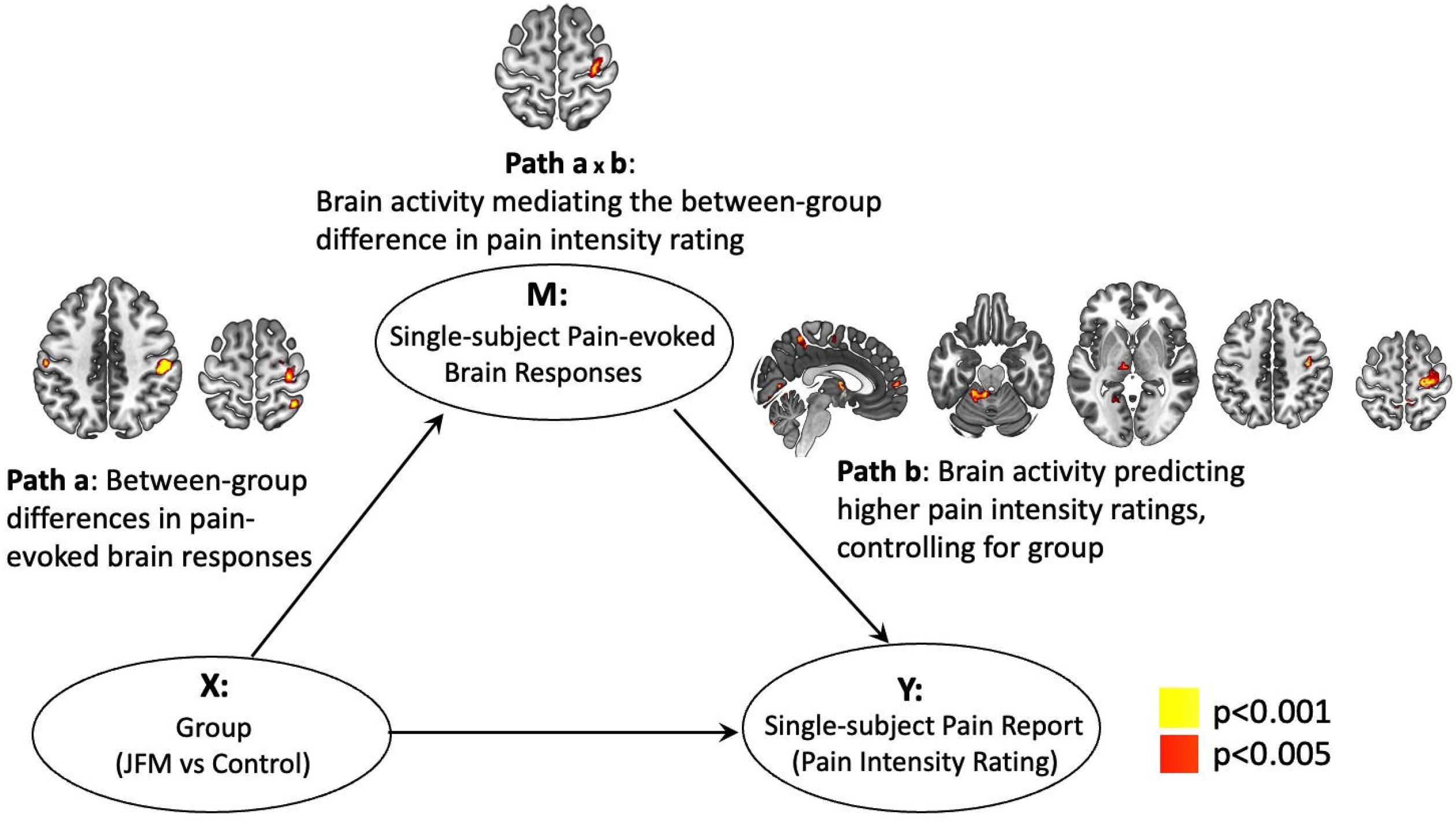
The model and results of exploratory whole-brain mediation analysis, which examines the relationships between group (X), single-subject pain-evoked brain responses to stimuli at 4 kg/cm^2^ (M), and single-subject pain intensity ratings to stimuli at 4 kg/cm^2^(Y). Pain-evoked brain responses in primary sensorimotor cortex mediates the between-group differences in pain intensity ratings.

## Discussion

This is the first study that directly compares pain-evoked brain responses between adolescents with JFM and healthy adolescents using a relatively large sample size. We found that, compared with healthy adolescents, adolescents with JFM were more sensitive to noxious pressure at both low (2.5 kg/cm^2^) and medium (4 kg/cm^2^) stimulus intensities and exhibited significantly augmented pain-related activation in the somatomotor network, particularly in the primary somatosensory cortex, whose activation correlated with greater widespreadness of clinical pain. We also found that amplified primary sensorimotor cortex activation in adolescents with JFM mediated between-group differences in pain ratings. Taken together, these findings suggest that adolescents with JFM have greater pain sensitivity and greater sensorimotor processing, which may reflect augmented peripheral nociceptive input to the brain or sensitization of the somatosensory system at the central level.

First, we compared pain intensity and unpleasantness ratings between adolescents with JFM and healthy controls using mixed-design ANOVA. We found significant main effects of group and stimulus intensity but no interaction, suggesting that adolescents with JFM are similarly hypersensitive to noxious pressure stimuli at low and medium intensities. This finding is in line with previous observations by our group in the 2017 King and colleagues’ study(16). In that study, mechanical pressure was applied to the forehead and the palm using a hand-held algometer utilizing a method of limits (i.e., pressure pain thresholds), and adolescents with JFM rated higher pain intensity of the pressure stimulus than healthy adolescents. In the current study, suprathreshold pressure was applied using a computer-controlled pressure pain device to the thumbnail bed, one of the most distal parts of the body, as well as the most commonly used site in comparable imaging studies on adult fibromyalgia(9, 11, 12). Our finding not only confirmed that adolescents with JFM are hypersensitive to noxious pressure but also added experimental evidence to the widespreadness of this increased pain sensitivity. The rest of the study sought to understand the underlying brain mechanisms.

The massive univariate GLM analyses showed that adolescents with JFM exhibited greater pain-evoked activation in the right S1, contralateral to the side of stimulation. The S1 region is almost always activated during acute experimental pain and is associated with sensory-discriminative aspects of pain perception(6). Our finding is consistent with previous studies showing augmented S1 responses during pain in adult fibromyalgia(9), suggesting a potential common brain mechanism between juvenile and adult forms of fibromyalgia. S1 hyperactivation during pain in adolescents with JFM could reflect central sensitization or disinhibition. For example, compromised intracortical inhibition in S1 has been observed in adults with fibromyalgia using magnetoencephalography (42). A review of neuroimaging literature also revealed that experimental pain increased the functional connectivity between S1 and insula in adults with fibromyalgia but not in controls, and this strengthened S1-to-insula connectivity was positively correlated with temporal summation of pain(15). Another possibility is that S1 hyperactivation results from increased or amplified nociceptive input to the brain. For instance, a recent study found that about half of adolescents with JFM have skin biopsy findings suggesting small-fiber neuropathy(43), consistent with findings in adults with FM(44). Another study found that some adults with FM have silent nociceptors exhibiting hyperexcitability(45).

Interestingly, we also found that S1 activation positively correlated with the Widespread Pain Index score, but not the Symptom Severity Scale score. This finding suggests that S1 hyper-activation in adolescents with JFM may only account for the sensory-discriminative aspect of pain perception but not the overall severity of symptoms in JFM.

Moreover, we demonstrated that adolescents with JFM had overall amplified responses in somatomotor network when experiencing low pain. The somatomotor network, also known as sensorimotor network, is a large-scale brain network that mainly includes primary somatosensory (S1, postcentral gyrus) and primary motor (M1, precentral gyrus) regions and extends to supplementary motor areas(46). The sensorimotor network is typically activated during motor tasks such as finger tapping(47) but also becomes activated during pain(6). In addition, randomized sham-controlled clinical trials have found that high-frequency repetitive transcranial magnetic stimulation targeting primary motor cortex (M1) significantly reduced clincal pain in adults with fibromyalgia(48, 49), further confirming the importance of the somatomotor network in pain perception and analgesia.

Finally, using whole-brain mediation analysis, we found that brain activity in primary sensorimotor cortex mediated the between-group difference in pain intensity ratings, which implies that sensorimotor responses in adolescents with JFM may account for their greater pain sensitivity compared to healthy adolescents.

This study has several limitations that should be considered when interpreting the results. First, only female adolescents, who represent the majority of patients with JFM, were included in our study. Therefore, our findings might not be applied to male adolescents and children with JFM. Although quite rare, they could have distinct characteristics of pain processing in the brain and warrrant further research. Secondly, the noxious pressure stimuli used in our study only elicited mild and moderate pain. We do not know whether there is difference in brain processing of severe pain between adolescents with JFM and their healthy peers. Thirdly, medication use in patients with JFM could have acted as a confounding factor, although all participants were required to maintain a stable regimen for at least three weeks before their fMRI scan. Lastly, our study is cross-sectional and included 33 participants per group. Future research is needed to reproduce our findings in larger samples and investigate whether these observed functional alterations in the brain respond to treatment.

In conclusion, this study provides the first evidence of greater pain-evoked brain responses in adolescents with JFM involving primary sensorimotor regions important for sensory-discriminative aspect of pain perception, which may account for their augmented pain sensitivity compared to healthy adolescents.

## Data Availability

All data produced in the present study are available upon reasonable request to the authors

## Acknowledgements

The authors gratefully thank Matt Lanier, Kaley Ireland, Kelsey Murphy, Brynne Williams, Sarah Miozzi and Lacey Haas (Imaging Research Center, Department of Radiology, Cincinnati Children’s Hospital Medical Center) for their assistance in collecting MRI data.

## References

1. Buskila D, Press J, Gedalia A, Klein M, Neumann L, Boehm R, et al. Assessment of nonarticular tenderness and prevalence of fibromyalgia in children. J Rheumatol. 1993;20(2):368–70.

2. Kashikar-Zuck S, Ting TV. Juvenile fibromyalgia: current status of research and future developments. Nat Rev Rheumatol. 2014;10(2):89–96.

3. Kashikar-Zuck S, Cunningham N, Peugh J, Black WR, Nelson S, Lynch-Jordan AM, et al. Long-term outcomes of adolescents with juvenile-onset fibromyalgia into adulthood and impact of depressive symptoms on functioning over time. Pain. 2019;160(2):433–41.

4. Ting TV, Barnett K, Lynch-Jordan A, Whitacre C, Henrickson M, Kashikar-Zuck S. 2010 American College of Rheumatology Adult Fibromyalgia Criteria for Use in an Adolescent Female Population with Juvenile Fibromyalgia. J Pediatr. 2016;169:181–7 e1.

5. Ting TV, Hashkes PJ, Schikler K, Desai AM, Spalding S, Kashikar-Zuck S. The role of benign joint hypermobility in the pain experience in Juvenile Fibromyalgia: an observational study. Pediatr Rheumatol Online J. 2012;10(1):16.

6. Apkarian AV, Bushnell MC, Treede RD, Zubieta JK. Human brain mechanisms of pain perception and regulation in health and disease. Eur J Pain. 2005;9(4):463–84.

7. Casey BJ, Getz S, Galvan A. The adolescent brain. Dev Rev. 2008;28(1):62–77.

8. Tong H, Maloney TC, Payne MF, King CD, Ting TV, Kashikar-Zuck S, et al. Processing of pain by the developing brain: evidence of differences between adolescent and adult females. Pain. 2022.

9. Gracely RH, Petzke F, Wolf JM, Clauw DJ. Functional magnetic resonance imaging evidence of augmented pain processing in fibromyalgia. Arthritis Rheum. 2002;46(5):1333–43.

10. Cook DB, Lange G, Ciccone DS, Liu WC, Steffener J, Natelson BH. Functional imaging of pain in patients with primary fibromyalgia. J Rheumatol. 2004;31(2):364–78.

11. Pujol J, Lopez-Sola M, Ortiz H, Vilanova JC, Harrison BJ, Yucel M, et al. Mapping brain response to pain in fibromyalgia patients using temporal analysis of FMRI. PLoS One. 2009;4(4):e5224.

12. Lopez-Sola M, Woo CW, Pujol J, Deus J, Harrison BJ, Monfort J, et al. Towards a neurophysiological signature for fibromyalgia. Pain. 2017;158(1):34–47.

13. Lopez-Sola M, Pujol J, Wager TD, Garcia-Fontanals A, Blanco-Hinojo L, Garcia-Blanco S, et al. Altered functional magnetic resonance imaging responses to nonpainful sensory stimulation in fibromyalgia patients. Arthritis Rheumatol. 2014;66(11):3200–9.

14. Pujol J, Macia D, Garcia-Fontanals A, Blanco-Hinojo L, Lopez-Sola M, Garcia-Blanco S, et al. The contribution of sensory system functional connectivity reduction to clinical pain in fibromyalgia. Pain. 2014;155(8):1492–503.

15. Kim J, Loggia ML, Cahalan CM, Harris RE, Beissner FDPN, Garcia RG, et al. The somatosensory link in fibromyalgia: functional connectivity of the primary somatosensory cortex is altered by sustained pain and is associated with clinical/autonomic dysfunction. Arthritis Rheumatol. 2015;67(5):1395–405.

16. King CD, Jastrowski Mano KE, Barnett KA, Pfeiffer M, Ting TV, Kashikar-Zuck S. Pressure Pain Threshold and Anxiety in Adolescent Females With and Without Juvenile Fibromyalgia: A Pilot Study. Clin J Pain. 2017;33(7):620–6.

17. Sunol M, Payne MF, Tong H, Maloney TC, Ting TV, Kashikar-Zuck S, et al. Brain Structural Changes during Juvenile Fibromyalgia: Relationships with Pain, Fatigue and Functional Disability. Arthritis Rheumatol. 2022.

18. Molina J, Amaro E, Jr., da Rocha LGS, Jorge L, Santos FH, Len CA. Functional resonance magnetic imaging (fMRI) in adolescents with idiopathic musculoskeletal pain: a paradigm of experimental pain. Pediatr Rheumatol Online J. 2017;15(1):81.

19. Wolfe F, Clauw DJ, Fitzcharles MA, Goldenberg DL, Katz RS, Mease P, et al. The American College of Rheumatology preliminary diagnostic criteria for fibromyalgia and measurement of symptom severity. Arthritis Care Res (Hoboken). 2010;62(5):600–10.

20. Daffin M, Gibler RC, Kashikar-Zuck S. Measures of Juvenile Fibromyalgia. Arthritis Care Res (Hoboken). 2020;72 Suppl 10:171–82.

21. Lopez-Sola M, Pujol J, Hernandez-Ribas R, Harrison BJ, Ortiz H, Soriano-Mas C, et al. Dynamic assessment of the right lateral frontal cortex response to painful stimulation. Neuroimage. 2010;50(3):1177–87.

22. Price DD, McGrath PA, Rafii A, Buckingham B. The validation of visual analogue scales as ratio scale measures for chronic and experimental pain. Pain. 1983;17(1):45–56.

23. Pruessmann KP, Weiger M, Scheidegger MB, Boesiger P. SENSE: sensitivity encoding for fast MRI. Magn Reson Med. 1999;42(5):952–62.

24. Smith SM, Jenkinson M, Woolrich MW, Beckmann CF, Behrens TE, Johansen-Berg H, et al. Advances in functional and structural MR image analysis and implementation as FSL. Neuroimage. 2004;23 Suppl 1:S208–19.

25. Cox RW. AFNI: software for analysis and visualization of functional magnetic resonance neuroimages. Comput Biomed Res. 1996;29(3):162–73.

26. Smith SM. Fast robust automated brain extraction. Hum Brain Mapp. 2002;17(3):143–55.

27. Zhang Y, Brady M, Smith S. Segmentation of brain MR images through a hidden Markov random field model and the expectation-maximization algorithm. IEEE Trans Med Imaging. 2001;20(1):45–57.

28. Fonov V, Evans AC, Botteron K, Almli CR, McKinstry RC, Collins DL, et al. Unbiased average age-appropriate atlases for pediatric studies. Neuroimage. 2011;54(1):313–27.

29. Jenkinson M, Smith S. A global optimisation method for robust affine registration of brain images. Med Image Anal. 2001;5(2):143–56.

30. Jenkinson M, Bannister P, Brady M, Smith S. Improved optimization for the robust and accurate linear registration and motion correction of brain images. Neuroimage. 2002;17(2):825–41.

31. Power JD, Barnes KA, Snyder AZ, Schlaggar BL, Petersen SE. Spurious but systematic correlations in functional connectivity MRI networks arise from subject motion. Neuroimage. 2012;59(3):2142–54.

32. Coghill RC, Sang CN, Berman KF, Bennett GJ, Iadarola MJ. Global cerebral blood flow decreases during pain. J Cereb Blood Flow Metab. 1998;18(2):141–7.

33. Woolrich MW, Ripley BD, Brady M, Smith SM. Temporal autocorrelation in univariate linear modeling of FMRI data. Neuroimage. 2001;14(6):1370–86.

34. Woolrich MW, Behrens TE, Beckmann CF, Jenkinson M, Smith SM. Multilevel linear modelling for FMRI group analysis using Bayesian inference. Neuroimage. 2004;21(4):1732–47.

35. Worsley KJ, Evans AC, Marrett S, Neelin P. A three-dimensional statistical analysis for CBF activation studies in human brain. J Cereb Blood Flow Metab. 1992;12(6):900–18.

36. Harris CR, Millman KJ, van der Walt SJ, Gommers R, Virtanen P, Cournapeau D, et al. Array programming with NumPy. Nature. 2020;585(7825):357–62.

37. Yeo BT, Krienen FM, Sepulcre J, Sabuncu MR, Lashkari D, Hollinshead M, et al. The organization of the human cerebral cortex estimated by intrinsic functional connectivity. J Neurophysiol. 2011;106(3):1125–65.

38. Wager TD, Davidson ML, Hughes BL, Lindquist MA, Ochsner KN. Prefrontal-subcortical pathways mediating successful emotion regulation. Neuron. 2008;59(6):1037–50.

39. Atlas LY, Bolger N, Lindquist MA, Wager TD. Brain mediators of predictive cue effects on perceived pain. J Neurosci. 2010;30(39):12964–77.

40. Koban L, Kross E, Woo CW, Ruzic L, Wager TD. Frontal-Brainstem Pathways Mediating Placebo Effects on Social Rejection. J Neurosci. 2017;37(13):3621–31.

41. Lopez-Sola M, Geuter S, Koban L, Coan JA, Wager TD. Brain mechanisms of social touch-induced analgesia in females. Pain. 2019;160(9):2072–85.

42. Lim M, Roosink M, Kim JS, Kim DJ, Kim HW, Lee EB, et al. Disinhibition of the primary somatosensory cortex in patients with fibromyalgia. Pain. 2015;156(4):666–74.

43. Boneparth A, Chen S, Horton DB, Moorthy LN, Farquhar I, Downs HM, et al. Epidermal Neurite Density in Skin Biopsies From Patients With Juvenile Fibromyalgia. J Rheumatol. 2021;48(4):575–8.

44. Oaklander AL, Herzog ZD, Downs HM, Klein MM. Objective evidence that small-fiber polyneuropathy underlies some illnesses currently labeled as fibromyalgia. Pain. 2013;154(11):2310–6.

45. Serra J, Collado A, Sola R, Antonelli F, Torres X, Salgueiro M, et al. Hyperexcitable C nociceptors in fibromyalgia. Ann Neurol. 2014;75(2):196–208.

46. Chenji S, Jha S, Lee D, Brown M, Seres P, Mah D, et al. Investigating Default Mode and Sensorimotor Network Connectivity in Amyotrophic Lateral Sclerosis. PLoS One. 2016;11(6):e0157443.

47. Biswal B, Yetkin FZ, Haughton VM, Hyde JS. Functional connectivity in the motor cortex of resting human brain using echo-planar MRI. Magn Reson Med. 1995;34(4):537–41.

48. Passard A, Attal N, Benadhira R, Brasseur L, Saba G, Sichere P, et al. Effects of unilateral repetitive transcranial magnetic stimulation of the motor cortex on chronic widespread pain in fibromyalgia. Brain. 2007;130(Pt 10):2661–70.

49. Mhalla A, Baudic S, de Andrade DC, Gautron M, Perrot S, Teixeira MJ, et al. Long-term maintenance of the analgesic effects of transcranial magnetic stimulation in fibromyalgia. Pain. 2011;152(7):1478–85.

