## Supplementary Material for "Augmented Pain-Evoked Primary Sensorimotor Cortex Activation in Adolescent Girls with Juvenile Fibromyalgia"

#### Supplementary Methods:

Inclusion and exclusion criteria for study participants

#### Supplementary Figures:

Figure S1. Noxious pressure stimulation task.

Figure S2. Intensity and extensiveness of bodily pain ratings

#### Supplementary Tables

Table S1. Brain responses to 2.5 kg/cm$^2$ stimuli in the JFM group.

Table S2. Brain responses to 2.5 kg/cm$^2$ stimuli in the control group.

Table S3. Brain responses to 4 kg/cm$^2$ stimuli in the JFM group.

Table S4. Brain responses to 4 kg/cm$^2$ stimuli in the control group.

Table S5. Comparison of brain responses to 4 kg/cm$^2$ between the JFM group and the control group.

Table S6. Comparison of brain responses to 2.5 kg/cm$^2$ adjusted for age between the JFM group and the control group.

Table S7. Comparison of brain responses to 4 kg/cm$^2$ adjusted for age between the JFM group and the control group.

Table S8. Pain-evoked brain responses within seven brain networks.

Table S9. Between-group comparison of Pain-evoked brain responses within brain networks.

### Supplementary Methods:

#### Inclusion and exclusion criteria for study participants

Inclusion criteria for the JFM group included: (1) having received a clinical diagnosis of JFM from a pediatric rheumatologist or a pain physician following the 2010 American College of Rheumatology (ACR) diagnostic criteria for fibromyalgia(1, 2); (2) reporting 5 or more tender points on the study day; (3) reporting an average pain intensity during the past week of at least 3 out of 10; and (4) reporting a Functional Disability Inventory (FDI) score of 7 or higher out of 60, indicating at least mild disability(3). Inclusion criteria for the control group included (1) being healthy both physically and psychologically (i.e., not diagnosed with chronic pain, psychiatric, neurological, or inflammatory disorders), (2) reporting an average pain intensity of 0-2 out of 10 for the past week. (3) reporting an FDI score below 7, indicating no disability. Those participants with a contraindication to magnetic resonance imaging (MRI) scanning, developmental delay, major neurological or psychiatric disorders, a positive pregnancy test, or taking opioid or psychotropic medication were excluded from the study. All participants included in the study were either not taking any medication or were under a stable medication regimen for a minimum of 3 weeks prior to the first MRI assessment.

1. Wolfe F, Clauw DJ, Fitzcharles MA, Goldenberg DL, Katz RS, Mease P, et al. The American College of Rheumatology preliminary diagnostic criteria for fibromyalgia and measurement of symptom severity. Arthritis Care Res (Hoboken). 2010;62(5):600-10.
2. Ting TV, Barnett K, Lynch-Jordan A, Whitacre C, Henrickson M, Kashikar-Zuck S. 2010 American College of Rheumatology Adult Fibromyalgia Criteria for Use in an Adolescent Female Population with Juvenile Fibromyalgia. J Pediatr. 2016;169:181-7 e1.
3. Walker LS, Greene JW. The functional disability inventory: measuring a neglected dimension of child health status. J Pediatr Psychol. 1991;16(1):39-58.

### Supplementary Figures

**Figure S1.** Noxious pressure stimulation task. (A) Details and timing of one stimulation cycle. (B) Illustration of the computerized visual analogue scales used in the study.

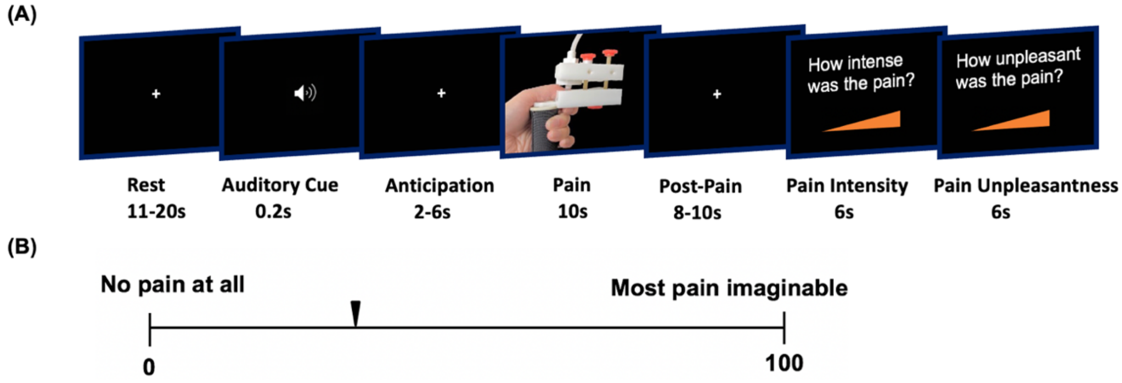


**Figure S2.** Intensity (A) and extensiveness (B) of bodily pain in healthy adolescents and adolescents with JFM. *** p<0.001 in two-sample t-test.

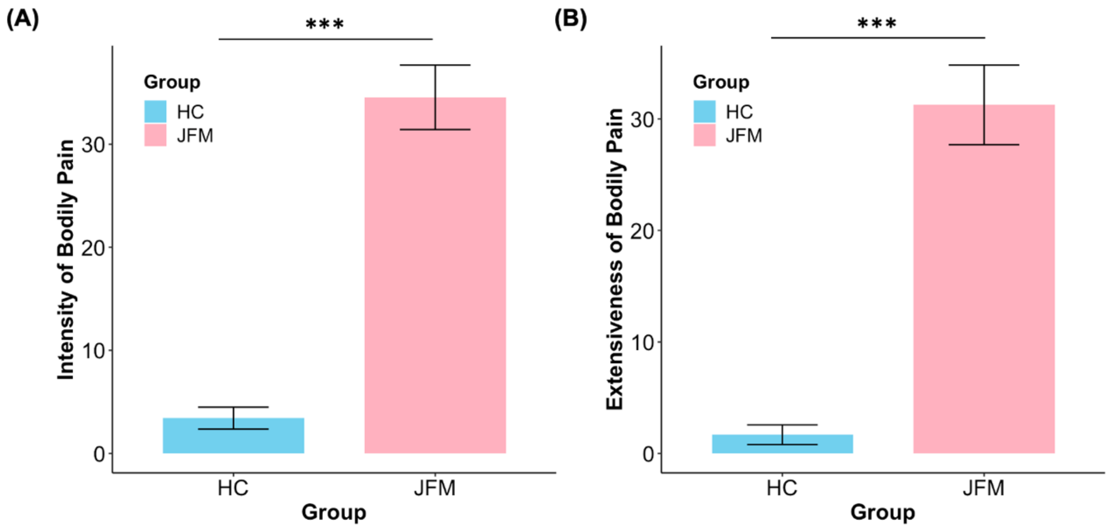

### Supplementary Tables

**Table S1.** Brain responses to 2.5 kg/cm$^2$ stimuli in the JFM group ($Z>3.1$, $p<0.05$, cluster-corrected).

| **Activation** | | | | | |
|---|---|---|---|---|---|
| **Cluster 1** | **Voxel Size:** 11135, **P-value:** 7.41E-11 | | | | |
| **Region** | **Side** | **X** | **Y** | **Z** | **Z-Score** |
| Parietal Operculum (S2) | L | -60 | -34 | 22 | 7.86 |
| Insular cortex | L | -40 | -6 | -10 | 7.75 |
| Supramarginal gyrus | L | -64 | -26 | 26 | 7.62 |
| Inferior frontal gyrus (DLPFC) | L | -44 | 36 | 8 | 5.67 |
| Precentral gyrus (M1) | L | -54 | 8 | 17 | 5.58 |
| Middle temporal gyrus | L | -58 | -66 | 8 | 4.56 |
| Putamen | L | -24 | 6 | -8 | 4.5 |
| Superior temporal gyrus | L | -64 | -30 | 8 | 4.39 |
| Postcentral gyrus (S1) | L | -58 | -14 | 48 | 4.1 |
| Middle frontal gyrus (Frontal pole) | L | -26 | 56 | 18 | 3.86 |
| Temporal pole | L | -32 | 8 | -28 | 3.46 |
| Thalamus | L | -2 | -15 | 0 | 3.43 |

| **Cluster 2** | **Voxel Size:** 26091, **P-value:** 8.85E-19 | | | | |
|---|---|---|---|---|---|
| **Region** | **Side** | **X** | **Y** | **Z** | **Z-Score** |
| Insular cortex | R | 40 | -12 | -6 | 7.85 |
| Parietal Operculum (S2) | R | 64 | -20 | 18 | 7.54 |

| | | | | | |
|---|---|---|---|---|---|
| Precentral gyrus (M1) | R | 58 | 10 | 0 | 7.45 |
| Frontopolar cortex | R | 46 | 40 | -2 | 7 |
| Supramarginal gyrus | R | 66 | -20 | 32 | 6.93 |
| Superior temporal gyrus | R | 62 | -32 | 20 | 6.91 |
| Postcentral gyrus (S1) | R | 58 | -16 | 40 | 6.74 |
| Anterior cingulate cortex | R | 4 | 8 | 32 | 6.32 |
| Anterior cingulate cortex | L | -6 | 22 | 28 | 5.84 |
| Inferior frontal gyrus (VLPFC) | R | 44 | 30 | -4 | 6.22 |
| Middle frontal gyrus (DLFPC) | R | 42 | 44 | 16 | 6.12 |
| Amygdala | R | 30 | -1 | -10 | 5.78 |
| Caudate | R | 13 | 14 | 2 | 4.1 |
| Caudate | L | -9 | 2 | 12 | 4 |
| Supplementary motor area | R | 2 | 4 | 56 | 5.56 |
| Supplementary motor area | L | -1 | 6 | 54 | 4.78 |
| Medial superior frontal gyrus (DMPFC) | R | 4 | 34 | 46 | 4.9 |
| Medial superior frontal gyrus (DMPFC) | L | -1 | 50 | 30 | 4.08 |
| Middle temporal gyrus | R | 50 | -22 | -11 | 3.95 |
| Thalamus | R | 8 | -10 | 10 | 3.89 |
| **Deactivation** | | | | | |
| **Cluster 1** | **Voxel Size:** 26142, **P-value:** 8.38E-19 | | | | |

| **Region** | **Side** | **X** | **Y** | **Z** | **Z-Score** |
|---|---|---|---|---|---|
| Calcarine cortex | R | 8 | -70 | 8 | 6.61 |
| | L | -2 | -70 | 8 | 5.59 |
| Precuneus | R | 6 | -68 | 58 | 6.29 |
| | L | -3 | -8 | 60 | 5.68 |
| Cerebellum | R | 20 | -54 | -52 | 6.26 |
| | L | -26 | -40 | -23 | 4.65 |
| Fusiform gyrus | R | 30 | -36 | -23 | 5.28 |
| | L | -32 | -40 | -23 | 4.38 |
| Hippocampus | R | -32 | -28 | -13 | 4.92 |
| | L | 33 | -25 | -13 | 3.92 |
| Lingual gyrus | R | 6 | -54 | 2 | 6.15 |
| | L | -6 | -60 | 9 | 5.17 |
| Middle occipital gyrus | L | 33 | -80 | 40 | 5.6 |
| | R | -30 | -82 | 40 | 5.48 |
| Thalamus | L | -18 | -30 | 7 | 4.89 |
| | R | 19 | -30 | 7 | 3.47 |
| Precentral gyrus | L | -37 | -10 | 64 | 4.24 |
| Inferior temporal cortex | R | 53 | -52 | -16 | 4.07 |
| | L | -52 | -52 | -16 | 3.85 |

**Table S2.** Brain responses to 2.5 $kg/cm^2$ stimuli in the control group ($Z>3.1$, $p<0.05$, cluster-corrected).

| **Activation** | | | | | |
|---|---|---|---|---|---|
| **Cluster 1** | **Voxel Size:** 39630, **P-value:** 8.65E-24 | | | | |
| **Region** | **Side** | **X** | **Y** | **Z** | **Z-Score** |
| Insula | R | 42 | 4 | -2 | 8.05 |
| | L | -39 | -2 | -4 | 6.54 |
| Postcentral gyrus (S1) | R | 60 | -16 | 18 | 7.67 |
| | L | -60 | -20 | 28 | 7.17 |
| Parietal operculum (S2) | R | 58 | -18 | 16 | 7.46 |
| | L | -62 | -22 | 17 | 6.47 |
| Supramarginal gyrus | R | 61 | -24 | 24 | 7.25 |
| | L | -54 | -26 | 20 | 7.02 |
| Superior temporal gyrus | R | 54 | -26 | 17 | 7.21 |
| | L | -60 | -23 | 14 | 5.64 |
| Amygdala | R | 27 | 5 | -14 | 6.18 |
| | L | -22 | 3 | -14 | 5.66 |
| Anterior cingulate cortex | R | 6 | 14 | 31 | 6.08 |
| | L | -5 | 36 | 16 | 5.13 |
| Precentral gyrus (M1) | R | 54 | 6 | 23 | 5.94 |
| | L | -54 | 8 | 15 | 5.18 |
| Caudate | R | 14 | 6 | 12 | 5.86 |
| | L | -15 | 6 | 12 | 4.59 |

| | | | | | |
|---|---|---|---|---|---|
| Putamen | R | 34 | 9 | -2 | 5.87 |
| | L | -31 | 10 | -2 | 5.26 |
| Thalamus | R | 5 | -13 | 17 | 5.59 |
| | L | -15 | -8 | 15 | 3.48 |
| Supplementary motor area | R | 6 | 18 | 52 | 5.8 |
| Parahippocampal gyrus | R | 22 | 7 | -20 | 5.72 |
| | L | -20 | 3 | -20 | 5.34 |
| Inferior frontal gyrus (VLPFC) | R | 47 | 40 | 2 | 6.37 |
| | L | -48 | 32 | 0 | 4.2 |
| Middle frontal gyrus (DLPFC) | R | 38 | 48 | 19 | 5.56 |
| | L | -36 | 43 | 23 | 5.62 |
| Middle temporal gyrus | R | 56 | -24 | -10 | 5.43 |
| Medial superior frontal gyrus (DMPFC) | R | 6 | 49 | 30 | 5.17 |
| | L | -1 | 32 | 36 | 4.27 |
| Periaqueductal gray | - | 3 | -30 | -5 | 3.84 |

| **Deactivation** | | | | | |
|---|---|---|---|---|---|
| **Cluster 1** | **Voxel Size:** 27290, **P-value:** 1.15E-16 | | | | |
| **Region** | **Side** | **X** | **Y** | **Z** | **Z-Score** |
| Cerebellum | R | 14 | -54 | -20 | 6.58 |
| | L | -38 | -48 | -26 | 5.51 |
| Fusiform gyrus | R | 28 | -37 | -21 | 6.35 |

| | | | | | |
|---|---|---|---|---|---|
| | L | -33 | -40 | -15 | 6.52 |
| Inferior temporal gyrus | L | -48 | -56 | -16 | 6.18 |
| | R | 50 | -55 | -16 | 4.44 |
| Precentral gyrus | L | -34 | 16 | 68 | 5.92 |
| Calcarine cortex | R | 8 | -56 | 12 | 5.91 |
| | L | -10 | -56 | 8 | 5.56 |
| Middle occipital gyrus | L | -36 | -77 | 29 | 5.82 |
| | R | 36 | -83 | 30 | 5.41 |
| Precuneus | L | -7 | -69 | 62 | 5.54 |
| | R | 7 | -68 | 60 | 5.33 |
| Hippocampus | L | -22 | -36 | 2 | 5.38 |
| | R | 18 | -34 | 2 | 4.09 |
| Brainstem (Pons) | L | -12 | -19 | -33 | 4.86 |
| Posterior Cingulate | R | 11 | -44 | 31 | 3.68 |

**Table S3.** Brain responses to 4 $kg/cm^2$ stimuli in the JFM group ($Z>3.1$, $p<0.05$, cluster-corrected).

| **Activation** | | | | | |
|---|---|---|---|---|---|
| **Cluster 1** | **Voxel Size:** 28807, **P-value:** 1.16E-24 | | | | |
| **Region** | **Side** | **X** | **Y** | **Z** | **Z-Score** |
| Insula | R | 42 | 6 | -12 | 8.02 |
| | L | -40 | -12 | 0 | 7.4 |

| | | | | | |
|---|---|---|---|---|---|
| Supramarginal gyrus | R | 66 | -22 | 34 | 7.17 |
| | L | -62 | -26 | 22 | 6.61 |
| Postcentral gyrus (S1) | R | 58 | -18 | 40 | 6.75 |
| | L | -60 | -20 | 27 | 6.43 |
| Superior temporal gyrus | R | 64 | -30 | 20 | 6.66 |
| | L | -60 | -32 | 20 | 6.28 |
| Parietal operculum (S2) | R | 54 | -32 | 24 | 6.45 |
| | L | -52 | 24 | 18 | 6.45 |
| Anterior cingulate cortex | R | 4 | 20 | 34 | 6.21 |
| | L | -6 | 24 | 28 | 6.18 |
| Precentral gyrus (M1) | R | 44 | -8 | 54 | 6.08 |
| | L | -54 | 8 | 15 | 5.37 |
| Inferior frontal gyrus (VLPFC) | R | 60 | 16 | 18 | 6.01 |
| | L | -54 | 10 | 18 | 5.11 |
| Putamen | R | 34 | 6 | 3 | 6 |
| | L | -31 | 12 | 3 | 5.4 |
| Middle frontal gyrus (DLPFC) | R | 30 | 52 | 18 | 5.92 |
| Amygdala | R | 26 | 4 | -18 | 5.9 |
| | L | -20 | 0 | -17 | 4.47 |
| Supplementary motor area | R | 5 | 4 | 58 | 5.64 |
| | L | -1 | 7 | 52 | 4.95 |
| Caudate | R | 10 | 3 | 9 | 4.64 |
| | L | -9 | 5 | 7 | 3.44 |

| | | | | | |
|---|---|---|---|---|---|
| Thalamus | R | 12 | -10 | 5 | 4.38 |
| | L | -6 | -5 | 9 | 3.51 |
| Medial superior frontal gyrus (DMPFC) | R | 6 | 49 | 30 | 5.17 |
| | L | -1 | 32 | 36 | 4.27 |
| Periaqueductal gray | - | 3 | -26 | -2 | 3.71 |
| **Cluster 2** | **Voxel Size:** 1852**, P-value:** 0.000301 | | | | |
| **Region** | **Side** | **X** | **Y** | **Z** | **Z-Score** |
| Cerebellum | L | -20 | -76 | -48 | 5.55 |
| **Deactivation** | | | | | |
| **Cluster 1** | **Voxel Size:** 23181**, P-value:** 2.21E-21 | | | | |
| **Region** | **Side** | **X** | **Y** | **Z** | **Z-Score** |
| Precuneus | R | 8 | -56 | 18 | 6.88 |
| | L | -2 | -62 | 18 | 5.85 |
| Fusiform gyrus | R | 28 | -38 | -16 | 6.71 |
| | L | -30 | -40 | -20 | 5.64 |
| Middle occipital gyrus | R | 46 | -72 | 26 | 6.41 |
| | L | -38 | -76 | 32 | 6.18 |
| Hippocampus | L | -30 | -24 | -14 | 6.39 |
| | R | 34 | -26 | -12 | 4.43 |
| Thalamus | R | 22 | -34 | 2 | 6.09 |
| | L | -18 | -34 | 6 | 5.96 |

| | | | | | |
|---|---|---|---|---|---|
| Lingual gyrus | R | 16 | -48 | 2 | 5.76 |
| | L | -10 | -46 | 4 | 5.44 |
| Cerebellum | R | 33 | -42 | -27 | 5.64 |
| Posterior Cingulate | R | 10 | -44 | 38 | 5.34 |
| | L | -4 | -34 | 36 | 4.85 |
| Cuneus cortex | R | 2 | -74 | 24 | 5.3 |
| | L | -12 | -92 | 28 | 5.19 |
| Inferior temporal cortex | L | -52 | -46 | -14 | 4.59 |
| | R | 47 | -50 | -8 | 4.01 |

**Table S4.** Brain responses to 4 kg/cm$^2$ stimuli in the control group ($Z>3.1$, $p<0.05$, cluster-corrected).

| **Activation** | | | | | |
|---|---|---|---|---|---|
| **Cluster 1** | **Voxel Size:** 37440**, P-value:** 1.58E-25 | | | | |
| **Region** | **Side** | **X** | **Y** | **Z** | **Z-Score** |
| Insula | R | 40 | 10 | -4 | 8.5 |
| | L | -42 | 2 | -6 | 8.24 |
| Supramarginal gyrus | R | 62 | -24 | 24 | 7.74 |
| | L | -62 | -34 | 30 | 6.61 |
| Postcentral gyrus (S1) | R | 62 | -18 | 20 | 7.33 |
| | L | -66 | -20 | 18 | 7.22 |
| Parietal operculum (S2) | R | 54 | -26 | 20 | 7.24 |

| | | | | | |
|---|---|---|---|---|---|
| | L | -60 | -28 | 18 | 7.02 |
| Superior temporal gyrus | R | 58 | 4 | -2 | 7.19 |
| | L | -54 | 6 | -2 | 7.05 |
| Amygdala | R | 26 | 4 | -16 | 6.86 |
| | L | -23 | 1 | -14 | 5.31 |
| Supplementary motor area | R | 8 | 26 | 58 | 6.49 |
| | L | -1 | 18 | 50 | 4.18 |
| Anterior cingulate cortex | R | 8 | 24 | 36 | 6.3 |
| | L | -1 | 37 | 17 | 5.9 |
| Putamen | R | 32 | 6 | -1 | 6.32 |
| | L | -31 | 10 | -3 | 5.66 |
| Pallidum | R | 18 | 6 | 0 | 6.23 |
| | L | -14 | 10 | -3 | 4.92 |
| Caudate | R | 12 | 6 | 10 | 6.15 |
| | L | -10 | 6 | 7 | 4.81 |
| Medial superior frontal gyrus (DMPFC) | R | 5 | 34 | 50 | 5.82 |
| | L | -1 | 44 | 42 | 4.23 |
| Thalamus | R | 7 | -7 | 8 | 5.76 |
| | L | -10 | -12 | 3 | 4.21 |
| Middle frontal gyrus (DLPFC) | R | 28 | 54 | 20 | 5.59 |
| | L | -35 | 43 | 23 | 5.76 |
| Precentral gyrus (M1) | R | 48 | 6 | 36 | 5 |
| | L | -54 | 8 | 17 | 4.81 |

| | | | | | |
|---|---|---|---|---|---|
| Inferior frontal gyrus (VLPFC) | R | 48 | 36 | 2 | 5.71 |
| | L | -36 | 38 | 12 | 4.46 |
| Periaqueductal gray | - | 2 | -30 | -10 | 4.15 |

| **Cluster 2** | **Voxel Size:** 1469, **P-value:** 0.00245 | | | | |
|---|---|---|---|---|---|
| **Region** | **Side** | **X** | **Y** | **Z** | **Z-Score** |
| Cerebellum | L | -20 | -70 | -54 | 5.98 |
| | R | 12 | -20 | -34 | 4.58 |
| **Deactivation** | | | | | |
| **Cluster 1** | **Voxel Size:** 30068, **P-value:** 4.08E-22 | | | | |
| **Region** | **Side** | **X** | **Y** | **Z** | **Z-Score** |
| Fusiform gyrus | L | -30 | -58 | -14 | 7.28 |
| | R | 28 | -40 | -20 | 6.82 |
| Middle occipital gyrus | L | -36 | -78 | 34 | 6.81 |
| | R | 22 | -80 | 42 | 6.56 |
| Lingual gyrus | R | 22 | -56 | 6 | 6.23 |
| | L | -20 | -48 | -5 | 5.62 |
| Calcarine cortex | R | 8 | -58 | 14 | 6.16 |
| | L | -9 | -60 | 8 | 4.42 |
| Cerebellum | R | 34 | -42 | -26 | 6.13 |
| | L | -28 | -36 | -25 | 5.03 |
| Precentral gyrus | L | -44 | -20 | 64 | 5.73 |
| Inferior temporal cortex | L | -48 | -54 | -17 | 5.66 |

| | | | | | |
|---|---|---|---|---|---|
| | R | 52 | -53 | -16 | 5.2 |
| Postcentral gyrus | L | -46 | -18 | 60 | 5.56 |
| Precuneus | R | 5 | -64 | 56 | 4.94 |
| | L | -13 | -65 | 54 | 4.94 |
| Hippocampus | L | -32 | -34 | -5 | 4.64 |
| | R | 32 | -33 | -2 | 3.46 |
| Thalamus | L | -19 | -29 | 14 | 3.36 |
| **Cluster 2** | **Voxel Size:** 1604**, P-value:** 0.00164 | | | | |
| **Region** | **Side** | **X** | **Y** | **Z** | **Z-Score** |
| Rectal gyrus | L | -8 | 46 | -20 | 5.28 |
| | R | 8 | 48 | -22 | 4.68 |

**Table S5.** Comparison of brain responses to 4 kg/cm$^2$ between the JFM group and the control group (Z>3.1, p<0.05, cluster-corrected).

| **Patient>Control** | | | | | |
|---|---|---|---|---|---|
| **Cluster 1** | **Voxel Size** | 288 | **P-value** | 0.00497 | |
| **Region** | **Side** | **X** | **Y** | **Z** | **Z-Score** |
| Postcentral gyrus | R | 48 | -26 | 46 | 4.76 |

**Table S6.** Comparison of brain responses to 2.5 $kg/cm^2$ adjusted for age between the JFM group and the control group ($Z>3.1$, $p<0.05$, cluster-corrected).

| **Patient>Control** | | | | | |
|---|---|---|---|---|---|
| **Cluster 1** | **Voxel Size** | 206 | **P-value** | 0.0155 | |
| **Region** | **Side** | **X** | **Y** | **Z** | **Z-Score** |
| Postcentral gyrus | R | 48 | -28 | 48 | 4.75 |

**Table S7.** Comparison of brain responses to 4 $kg/cm^2$ adjusted for age between the JFM group and the control group ($Z>3.1$, $p<0.05$, cluster-corrected).

| **Patient>Control** | | | | | |
|---|---|---|---|---|---|
| **Cluster 1** | **Voxel Size** | 262 | **P-value** | 0.00946 | |
| **Region** | **Side** | **X** | **Y** | **Z** | **Z-Score** |
| Postcentral gyrus | R | 48 | -26 | 46 | 4.88 |

**Table S8.** Pain-evoked brain responses within seven brain networks (results of one-sample t-tests).

| Stimulus Intensity | 2.5kg/cm$^2$ | | | | 4kg/cm$^2$ | | | |
|---|---|---|---|---|---|---|---|---|
| Group | JFM | | Control | | JFM | | Control | |
| Brain Network | t | p | t | p | t | p | t | p |
| Somatomotor | 8.3 | <0.001 | 3.34 | 0.002 | 6.4 | <0.001 | 2.94 | 0.006 |
| Dorsal Attention | -4.29 | <0.001 | -4.39 | <0.001 | -2.6 | 0.014 | -3.84 | <0.001 |
| Ventral Attention | 15.01 | <0.001 | 13.5 | <0.001 | 12.8 | <0.001 | 16.6 | <0.001 |
| Frontoparietal | 4.04 | <0.001 | 5.74 | <0.001 | 2.07 | 0.046 | 3.73 | <0.001 |
| Default Mode | 1.58 | 0.12 | 1.58 | 0.12 | -0.8 | 0.43 | 0.91 | 0.37 |
| Limbic | 0.01 | 0.99 | -0.72 | 0.48 | -1.97 | 0.058 | -0.05 | 0.96 |
| Visual | -8.45 | <0.001 | -6.12 | <0.001 | -6.96 | <0.001 | -8.02 | <0.001 |

**Table S9.** Between-group comparison of Pain-evoked brain responses within brain networks (results of two-sample t-tests).

| | JFM vs. Control | | | |
|---|---|---|---|---|
| Stimulus Intensity: | 2.5 kg/cm$^2$ | | 4 kg/cm$^2$ | |
| Brain Network | t | p | t | p |
| Somatomotor | 2.59 | 0.012 | 1.51 | 0.13 |
| Dorsal Attention | 0.15 | 0.88 | 0.96 | 0.34 |
| Ventral Attention | 0.75 | 0.46 | -0.43 | 0.67 |
| Frontoparietal | -0.12 | 0.91 | -0.33 | 0.74 |
| Default Mode | 0.16 | 0.88 | -1.2 | 0.23 |
| Limbic | 0.48 | 0.64 | -1.31 | 0.2 |
| Visual | -0.8 | 0.43 | 0.68 | 0.5 |
